# Identification of coxsackievirus A24 variant during an acute hemorrhagic conjunctivitis outbreak in coastal Kenya, 2024

**DOI:** 10.1101/2024.12.04.24318475

**Authors:** Arnold W. Lambisia, John Mwita Morobe, Edidah Moraa, Salim Mwarumba, Fredrick K.N. Korir, Raila Seif Athman, Rebecca Kiptui, Micheal Mbee, Nelly Mugo, Patrick Amoth, Penny Muange, Charlotte J. Houldcroft, Edwine Barasa, Joseph Mwangangi, George Githinji, Edward C. Holmes, Lynette Isabella Ochola-Oyier, Charles N. Agoti

## Abstract

**Background:** In early 2024, a surge in acute hemorrhagic conjunctivitis (AHC), also referred as “red eye” disease, was observed in coastal Kenya, prompting the Ministry of Health to issue an outbreak alert. Herein, we investigated the etiology of this outbreak.

**Methods:** Ocular swabs were obtained from 13 individuals presenting with AHC at a Mombasa clinic in early February 2024. Ten of these were analyzed using bacterial cultures, and all 13 using a pan-adenovirus quantitative PCR (qPCR) and metagenomic sequencing. Potential viral etiology was confirmed by a specific qPCR, amplicon sequencing and phylogenetic analysis.

**Results:** Bacterial cultures yielded no growth except in three samples where non-pathogenic bacteria were detected. All 13 samples were adenovirus qPCR negative.

Metagenomic sequencing detected coxsackievirus A24 variant (CA24v) in three of the 13 samples. CA24v detections were confirmed by both CA24v specific qPCR and amplicon sequencing of an approximately 450 nucleotide long VP4/2 junction genomic region. Phylogenetic analysis of the VP4/2 sequences showed that they were closely related to CA24v genotype IV.

**Conclusion:** The AHC epidemic in coastal Kenya in early 2024 was likely caused by CA24v. Metagenomic sequencing is a powerful tool for identifying potential causative agents of new disease outbreaks.

## Background

Acute haemorrhagic conjunctivitis (AHC) is a highly contagious eye disease first reported in 1969 in Ghana [1]. The syndrome, also referred to as “red eye”, “pink eye” or “Apollo 11 disease”, is characterised by one or more symptoms that include sudden onset ocular pain, foreign body sensation, sub-conjunctival haemorrhage, itching, photophobia, swollen eyelids, eye discharge, epiphora and blurred vision [2]. Systemic symptoms like fever, runny nose, congestion, limb pain, fatigue, and preauricular lymphadenopathy may also appear in some cases. Several AHC outbreaks have been reported in tropical and subtropical countries [3].

AHC disease is often self-limiting but results in individuals missing school or work, leading to economic losses [4], and school closure and/or case exclusion is a common strategy to curtail AHC transmission. During outbreaks, increased ophthalmologist clinic visits occur, and antibiotic prescription is common [5, 6]. The latter can indirectly contribute to the development of antibiotic resistance as most epidemics are in fact caused by viruses [7, 8]. Severe complications following infection are rare except for limited cases of radiculomyelitis (inflammation of the spinal root nerves and cord in ∼1 in 10,000 infections) [5, 9]. Mismanagement including self-medication and visiting traditional healers can exacerbate the situation [10, 11].

AHC can result from viral, bacterial or allergen exposure [12–14]. Viral causes are characterised by explosive outbreaks, with a short incubation time (12-48 hrs) and rapid spread [15–17]. Three major viruses are associated with AHC; specific adenovirus types, enterovirus 70, and the coxsackievirus A24 variant (CA24v) [5].

Other viruses such as SARS-CoV-2, measles, herpes simplex virus and varicella zoster virus have also been implicated in AHC clinical cases [14, 18]. Adenovirus AHC is mostly accompanied with keratitis [10, 19]. Laboratory diagnosis is therefore key to confirming AHC aetiology [20].

Following the COVID-19 pandemic, AHC outbreaks have been reported in Asia (India, Pakistan, Nepal) since 2022 [18, 21–24] and more recently in multiple countries in East and Southern Africa. In January 2024, the Kenya Ministry of Health issued an outbreak alert over increased cases viral conjunctivitis on the Kenyan Coast. The disease was reported to have spread extensively especially in Mombasa and Kilifi counties, with sporadic cases observed elsewhere in the country. We present the first investigation in Africa identifying CA24v as a potential etiological agent of 2024 AHC outbreaks in the region.

## Methods

### Ethical consideration

The samples analysed here were collected as part of Ministry of Health outbreak response activities to the AHC outbreak and as such written informed consent is not considered an essential step prior specimen collection. Molecular diagnostics and sequencing analysis in scenarios of outbreak response by Kenya Medical Research Institute (KEMRI) - Wellcome Trust Research Programme (KWTRP) has been approved by KEMRI Scientific Ethics Review Unit (SERU) Committee based in Nairobi, Kenya (Protocol #: **KEMRI/SERU/CGMR-C/304/4894**).

### Study population

On February 6^th^ 2024, 13 patients attending an outpatient eye clinic with unilateral or bilateral AHC, in Mombasa, provided an eye swab. No additional demographic details about the patients were shared. Mombasa city is Kenya’s second largest city, has a population of ∼ 1.2 million people, is located on the Kenyan coast and has a tropical climate. The samples were collected using a sterile general-purpose swab and immediately placed into universal transport media (UTM) and then transported to KWTRP, Kilifi, Kenya, at ambient temperature.

### Bacterial culture

To observe bacterial growth, the swabs from 10 samples were inoculated in blood agar, chocolate agar and MacConkey agar then incubated at 37°C in 5% CO_2_ for 18 hours. The remaining three lacked labels and were excluded from bacterial culture processing.

### Pan-adenovirus screening

Nucleic acids including DNA were extracted from 140µl of the 13 ocular swabs UTM by QIAamp Viral Mini-RNA kit (QIAGEN, UK) following the manufacturer’s instructions. These were then screened using a pan-adenovirus real-time PCR assay targeting the hexon gene of adenoviruses [25]. The following primer sequences were used: forward primer (5’ - GCCCCAGTGGTCTTACATGCACATC - 3’), probe (FAM - TCGGAGTACCTGAGCCCGGGTCTGGTGCA – MGBNFQ), and reverse primer (5’ – GCCACGGTGGGGTTTCTAAACTT - 3’). The thermocycling conditions were set up as follows: 95°C for 20 □ seconds and 40 cycles of 94°C for 15□seconds and 60°C for 30□seconds.

### Metagenomic screening

The nucleic acid extracts extracted above (n=13) were treated with TURBO™ DNase (Invitrogen, Carlsbad, CA) to deplete any DNA in the sample. First strand synthesis was performed using Superscript IV first strand synthesis kit and FR26RV-ENDOH primers as per the manufacturer’s instructions [26]. Second strand synthesis was done using Klenow fragment 3’ to 5’ exo- (New England BioLabs) and amplified using Q5® Hot Start High-Fidelity 2X Master Mix (New England BioLabs) and FR20RV primer (5’-GCCGGAGCTCTGCAGATATC-3’). The PCR product was used to prepare libraries using the SQK-LSK114 ligation kit with SQK-NBD114.96 native barcoding Kit and sequenced on the GridION Flow Cell (R10.4.1).

### Confirmation of detection of CA24v and VP4/VP2 sequencing

CA24v is a positive-sense RNA virus from the genus *Enterovirus*, family *Picornaviridae*. The single-stranded CA24v genome is approximately 7400 bp in length and encodes 4 capsid proteins (VP4, VP2, VP3, and VP1) and 7 non-structural proteins (2A-2C, and 3A-3D). Molecular epidemiological studies of CA24v often utilize the VP1 (receptor binding) or 3C (viral protease) regions of the CA24v genome. Currently, eight genotypes of CA24v are described (I–VIII) and responsible for major global epidemics of AHC [27].

To confirm the presence CA24v, all 13 samples were reanalysed using a CA24v specific quantitative PCR targeting the 5’ untranslated region (UTR) genomic region. Primers/probe sequences from a previous study were used [28]. The sequences were as follows: Forward primer: 5’-CCAACCACGGAGCAGGTGA-3’. Reverse primer: 5’-GAAACACGGACACCCAAAGTAGT-3’. Probe: 5’-CAACCCAGCAACTAGCCTGTCGTAACGC -3’. Additionally, the nucleic acid extracts from the positive samples were amplified using VP4/VP2 primers (Forward primer: CCGGCCCCTGAATGYGGCTAA and Reverse primer: TCWGGHARYTTCCAMCACCANCC) as previously described [29]. Library preparation was performed using the Illumina COVIDSEQ kit as per the manufacturer’s protocol, and the libraries were then sequenced on the Illumina Miseq platform.

### Data analysis

#### Metagenomic data

The barcodes and adapters were trimmed from the sequence data using porechop v.0.2.4. Taxonomic classification was then performed on the trimmed data to identify potential pathogens in the samples using the Nanopore mNGS Pipeline v.0.7 available on the Chan Zuckerberg ID webserver (i.e., CZ ID) [30].

#### Amplicon sequence data

The fastq data was checked for quality using FastQC v0.12.1, cleaned using FastP v0.23.4, mapped to the reference (Genbank accession PP548237) and the consensus genome generated using ivar v.1.3.1 [31].

#### Phylogenetic analysis

The VP4/VP2 segments of CA24v obtained through amplicon sequencing, as well as VP3 segments obtained from metagenomic sequencing, were used to estimate maximum likelihood trees. Global CA24v genomes were downloaded from GenBank on 05/07/2024. The recovered partial sequences were aligned using MAFFT v7.487, with the global CA24v sequences, trimmed to partial VP3 for the metagenomic data and VP4/VP2 segments for the amplicon sequencing data. Maximum likelihood trees were then estimated using the IQ-Tree2 v 2.1.3 software, employing the best-fit model of nucleotide substitution identified using the lowest Bayesian Information Criterion (BIC) and 1000 bootstrap replicates. Representative sequences from the eight known CA24v genotypes [27] were included on the phylogeny to identify the genotype present in the newly obtained Kenya CA24v sequences.

## Results

### Bacterial culture and adenovirus screening

No growth was observed in seven of the 10 processed samples through bacterial culture. Three cultures established non-pathogenic bacterial growth (*Actinocorallia libanotica*, *Lactobacillus plentarum,* and *Staphylococcus epidermidis*). Adenovirus quantitative PCR screening was negative for all 13 samples processed.

### Metagenomic sequencing and genotyping

All 13 samples processed by metagenomic sequencing. A total of 5.94 million reads were recovered, although most (∼90%) mapped to the human genome or non-pathogenic organisms (**Table 1**). None of the bacteria detected were likely pathogens. However, interestingly, in three samples, metagenomic sequence reads that mapped to CA24v were detected (Table1). In addition, three samples contained genetic sequence reads that mapped to rotavirus group A (RVA), and one had sequence reads that mapped to both CA24v and RVA. All the 13 samples were reanalysed using a CA24v and RVA specific quantitative PCR targeting the NSP3 genome segment [32]. Whereas all the samples gave a RVA negative PCR, the three samples with CA24v reads amplified with a cycle threshold values of 32.29, 38.43 and 37.65.

**Table 1.**
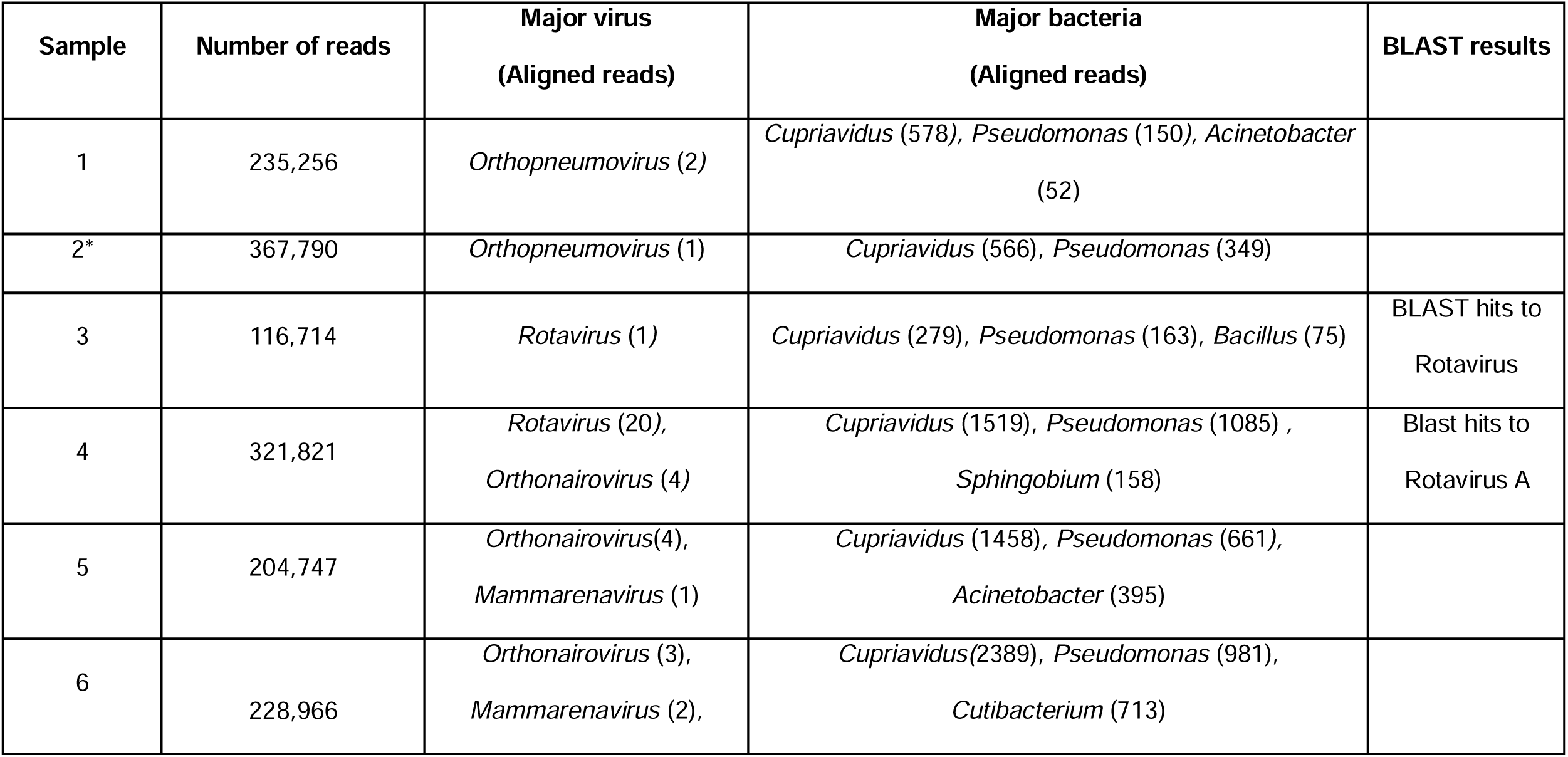
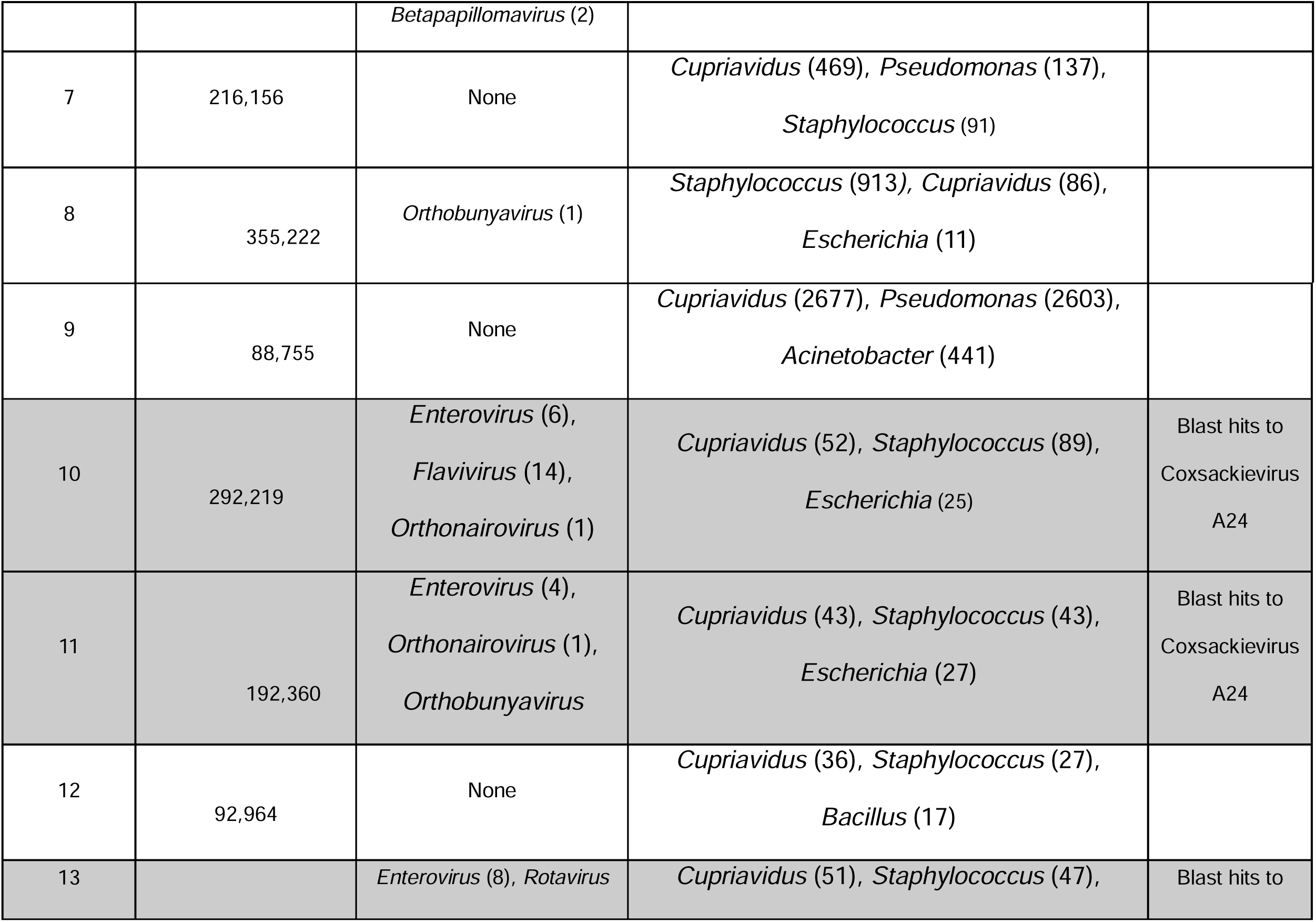
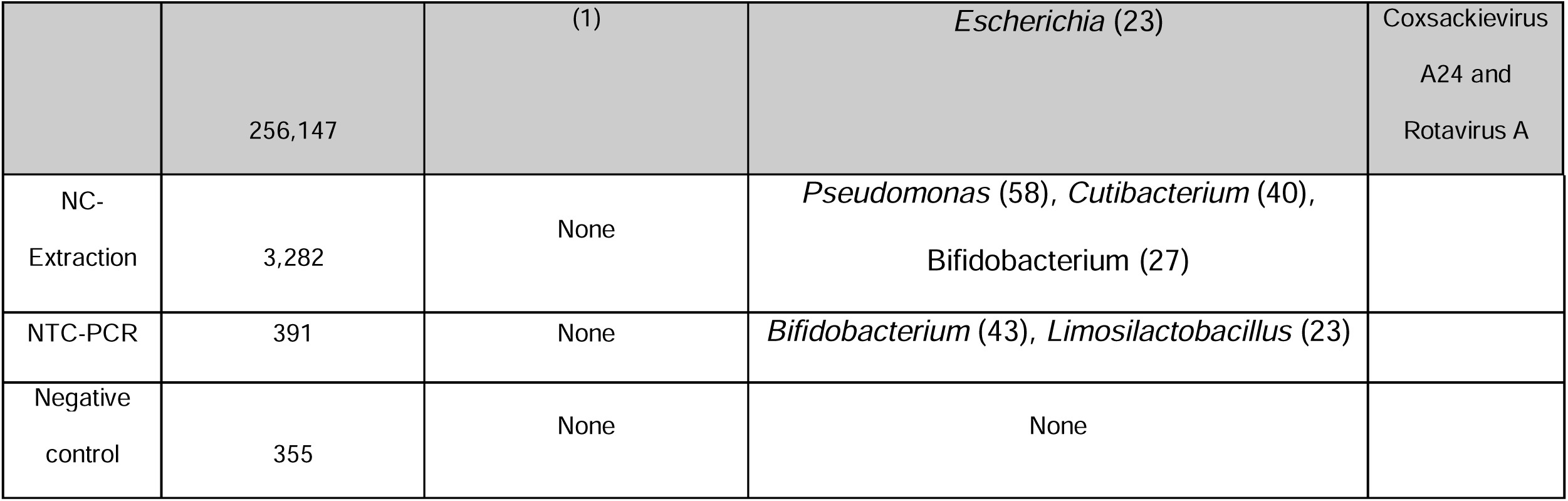
Results of the metagenomic analysis. A total of 13 clinical samples were processed together with an NC-Extraction, NTC-PCR, NC-library prep. Samples that contain CA24v are shaded grey

To characterize the strain of CA24v in the three coastal Kenya samples, we amplified and sequenced the VP4/2 junction region and undertook phylogenetic analysis including known genotype representative sequences. The alignment of the three Kenya CA24v VP4/2 sequences showed presence of 3 nucleotide changes. Phylogenic analysis showed that the sequences documented here clustered closely with the GIV genotype (**Figure 1**). Further, the coastal Kenya sequences showed a close relationship with sequences from Mayotte (Indian Ocean Island) that were sampled in 2024. Based on the VP1 sequences, the Mayotte sequences cluster within the cluster IV of the GIV genotype like Kenyan sequences sampled in 2010.

**Figure 1.**
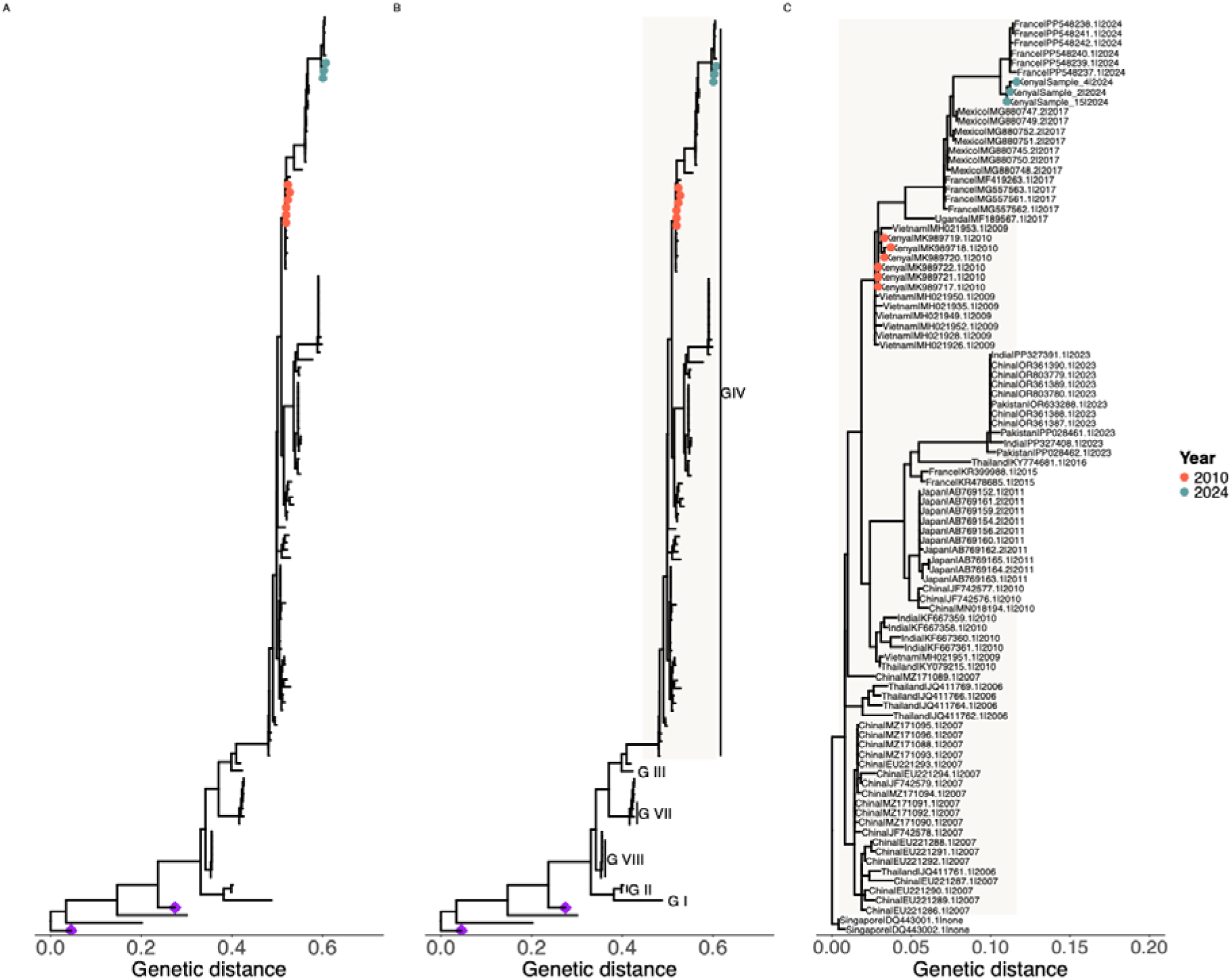
Phylogenetic placement of coxsackievirus A24 variant (CA24v) sequences from Kenya in a global context. *Panel A* shows the phylogeny based on VP3 sequences (n=132) including the Kenyan CA24v sequences recovered from metagenomic sequencing. *Panel B* shows the phylogeny based on VP4/VP2 sequences (n=158) including the Kenya data recovered by amplicon sequencing. *Panel C* shows a detailed phylogeny of genotype IV, showing the Kenyan sequences within the global context. The colored tip-points show Kenyan sequences from 2010 (orange) and 2024 (green). The purple diamonds show environmental CA24v sequences that clustered differently from the rest of the sequences.

## Discussion

Like in many parts of the world, the causes of AHC epidemics are not typically investigated in Kenya. This can be attributed to the limited diagnostic capacities in most outbreak settings and the self-limiting nature of the syndrome. However, several potential pathogens can be responsible, making it important to understand aetiology for appropriate public health advice. In the past, adenoviruses were mostly assumed to be the cause of AHC outbreaks, but this was rarely confirmed.

On 22^nd^ January 2024, the Kenya Ministry of Health issued an alert of an epidemic of AHC with the coastal region recording most cases [33]. We set out to determine disease etiology by processing a small set of samples using a pan-adenovirus quantitative PCR analysis and bacterial culture on ocular swabs from symptomatic cases. These returned negative results. However, by using metagenomic sequencing we detected CA24v sequences in three cases that were then confirmed by qPCR.

CA24v was first isolated in Singapore in 1970 [34], and has been recognized as one of the leading causes of AHC epidemics worldwide, typically occurring in densely populated tropical and subtropical areas. Few outbreaks of CA24v have occurred in the western hemisphere (mostly localised [35, 36]), likely due to better hygiene standards, limited overcrowding, and unfavourable climate conditions. Since 2022, there have been several reports of CA24v AHC outbreaks in parts of Asia and Africa [37–41].

To date, there have been few phylogenetic studies of CVA24v-related AHC outbreaks in the Africa [41]. Our metagenomic sequencing recovered part of the VP3 gene (135 bases) while amplicon sequencing generated parts of VP4/VP2 (450 bases), regions of the CA24v genome that are rarely used in CA24v genomic epidemiology. Phylogenetic analysis of these sequences in the global contextual data indicated the circulating CA24 strains in Kenya fell within genotype IV (GIV) that has been the dominant genotype in many recent outbreaks globally [41].

This is the first report of confirmed detection of CA24v in cases of AHC in the Kenyan population. We previously detected CA24v during a household-based respiratory syncytial virus (RSV) surveillance study in Kilifi, coastal Kenya, in 2010 [42]. Six cases (aged between 8.5-33 months) were identified from 95 metagenomically sequenced respiratory samples. The cases originated from six different households, none presented with conjunctivitis, one had diarrhoea, but all had rhinorrhoea. Based on VP1 sequences, the 2010 detections formed a monophyletic group within genotype GIV, but separate to that observed here for the 2024 outbreak.

The CA24v outbreak reported here is one of the multiple AHC outbreaks recorded in East Africa over the last two decades. CA24v was confirmed to be responsible for an AHC outbreak in June 2010 and July 2010, in Uganda (>26 districts, >6000 cases) and Southern Sudan (>400 cases), respectively [43]. Between November 2016 and January 2017, a CA24v AHC outbreak was observed in a prison, remand and police station in Gulu district, Uganda [44]. In 2024, AHC outbreaks have been reported in several countries in Eastern and Southern Africa including Tanzania [45], Uganda [46], Burundi [47], Malawi [48], Zambia [49], South Sudan [50] and South Africa [51]. In Kenya the epidemic seems to have spread beyond the coastal region to western region (Busia, Kisii, Siaya) and Nairobi City.

Our analysis here was limited by the small number of samples that were all collected from one geographic region in the country with no additional metadata. In addition, our metagenomic data comprised six million reads from a single GridION flow cell, predominantly mapping to the human genome, with few aligning to viruses as expected. Viral detections such as rotavirus that were not confirmed by targeted qPCR were ignored henceforth. The non-detection of CA24v in some samples may be attributed to (a) low sample quality (at collection, transport, storage), (b) timing of sample collection (i.e., the different stages of illness), and (c) the low sensitivity of metagenomic sequencing at this depth. Further, we cannot rule out that some of the AHC cases were a result of other causes than CA24v. As our metagenomic and amplicon sequencing recovered portions of CA24v genome that are not commonly used in genomic epidemiology, we are now establishing a whole genome sequencing platform to conduct further work on the virus.

This is the first study in Africa to identify CA24v as a possible cause of the recent AHC outbreaks in the region. Our findings were promptly shared with the Ministry of Health to assist with a public health response, and by collaborating with the Ministry we were able to determine the most likely cause of the AHC epidemic. Our study also demonstrates the power of metagenomic sequencing in identifying etiological agents in new epidemics. Despite this, several questions remain unanswered concerning CA24v epidemiological patterns and strain dynamics in the local population and the broader region. Detailed longitudinal investigations are needed to address these questions.

## Acknowledgements

We thank the laboratory members of the Pathogen Epidemiology and Omics (PEO) Group who support sample management and molecular diagnostics for outbreak samples. Funding for this analysis is from a Wellcome (grant no. 226002/A/22/Z), The Rockefeller Foundation (Grant OXF-FDG01), Cambridge-Africa ALBORADA Research Fund, and National Institute of Health and Care Research (grant. NIHR156467). This working is published with permission from director KEMRI.

## Data availability

The VP4/2 sequences have been deposited into GenBank under accession numbers (PQ034605, PQ034606 and PQ034607).

## Authors contributions

LIO-O, GG, CAN conceptualised the study

FK, MM, NM, PM oversaw sample collection

SM did the bacterial cultures

EM, JM, AWL did the metagenomic sequencing and phylogenetic analysis

AWL, JMM & CNA wrote the first draft of the manuscript.

ECH, CNA, LIO-O, GG, JM, PA, EB, CH, and NM, revised the manuscript.

LIO-O, GG, CNA provided grant funding and overall project supervision

